# PREVELANCE OF ISCHEMIC AND HEMORRAGHIC STROKE AMONG GERAITIC PATIENTS ADMITTED IN THE PUBLIC TERTIARY CARE HOSPITALS OF PESHAWAR

**DOI:** 10.1101/2023.03.30.23287970

**Authors:** Zeeshan Haider, Sayed Sajid Hussain

**Affiliations:** Sarhad University of Sciences and information Technology, Peshawar

**Keywords:** prevalence, Stroke, ischemic stroke, hemorrhagic stroke

## Abstract

**Background:** Stroke is a life-threatening medical condition that can result in lifelong brain impairment, complications, and demise. Stroke is the world’s second biggest cause of mortality and could soon overtake as the biggest cause of death globally. It has 2 major pathological types’ i.e. ischemic stroke and hemorrhagic stroke. Hypertension, diabetes mellitus, cardiac diseases, smoking, physical inactivity and age are the risk factors that contribute in the occurrence of a stroke.

**Objective:** To find out the epidemiological status of stroke types among patients admitted in the public tertiary care hospitals of Peshawar.

**Methodology:** A descriptive cross-sectional study was carried out to determine the prevalence of stroke types in tertiary care hospitals of Peshawar. The sample size calculated for the research study was 109. Convenience sampling technique was used in this study.

**Results:** This study was performed among 109 research participants. The most affected were males as 51.13% and females were 45.87%.And the rate of ischemic stroke were 71% while that of hemorrhagic stroke were 28%.

**Conclusion:** the current research study concluded that majority of the patients had ischemic stroke as compared to the hemorrhagic stroke.

## INTRODUCTION

Stroke is a life-threatening medical condition that can result in lifelong brain impairment, complications, and demise. Stroke is the world’s second biggest cause of mortality and could soon overtake as the biggest cause of death globally. A stroke is the loss of brain functions that occurs rapidly as a result of a disruption in the blood vessels providing blood to the brain. This can be produce by ischemia from a thrombosis or embolism, or a hemorrhage. As a result, the brain’s damaged area is unable to operate, causing one or more limbs on one side of the body to _become_ immobile, inability to recognize or produce speech, or seeing only one side of the visual field(Marwat, Usman, & Hussain, 2009).

According to Mumtaz Ali Marwat, et al. an observational study was conducted in Pakistan under the title Stroke and its relationship to risk factors. Total of 88 people were included in analysis. The mortality rate in stroke was 27.2% (Marwat, Usman, & Hussain, 2009)

According to Tapas Kumar Banerjee, et al. a population-based survey was conducted in different parts of India under the title Epidemiology of stroke in India that reviewed data from last decade of about 100,000 patients and the prevalence rate for stroke was 1.2%(Banerjee & Das, 2006).

Ischemic stroke happens when a blockage in the blood vessel of the neck or brain occurs, and is the most common type of stroke. This blockage can be produced by, “the development of a clot in a brain or neck blood artery, known as thrombosis; the migration of a blood clot from one section of the body to another such as the heart to the brain, known as embolism; or a serious narrowing of a cerebral artery, known as “stenosis” (Kummer et al., 2019).

In hemorrhagic stroke a blood vessel on surface of the brain ruptures which allow the blood to enter the gap between the skull and brain or when a faulty artery in the brain ruptures, causing blood to flood the surrounding tissue (Shah et al., 2013).

## METHODS

The study design was a descriptive cross-sectional study (from February 2022 to July 2022). After approval of Advance Studies and Research Board (ASRB), graduate committee and Ethical board of Sarhad University of Science and Information Technology the data was collected. The sample size calculated for this study was 109 through Raosoft. Non-probability Convenience sampling technique was used and data was analyzed by using software SPSS 25.

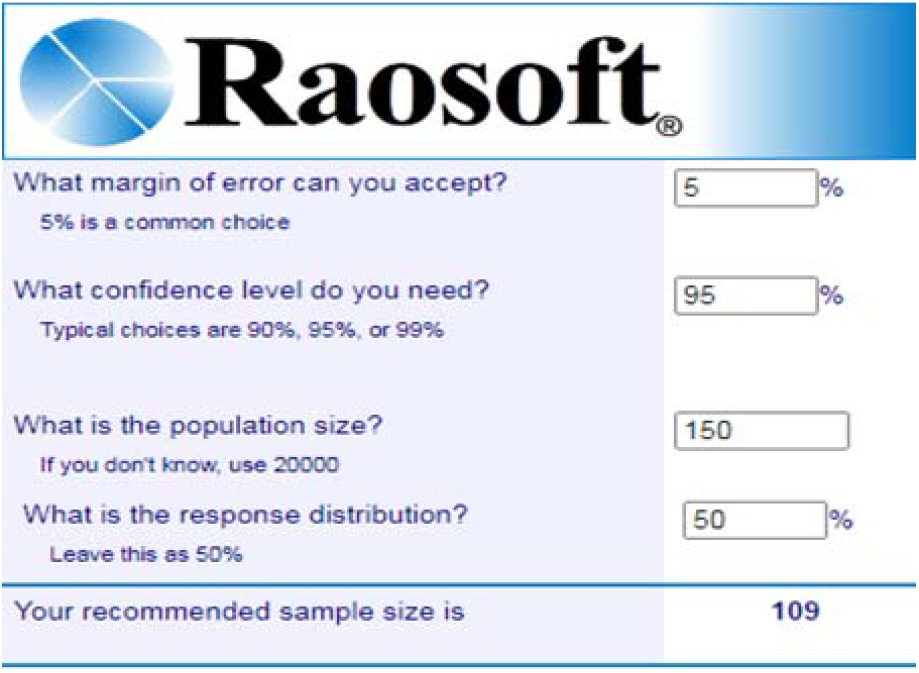

### Inclusion Criteria

- Those patients who are admitted in hospitals.
- Both male and female patients are included.
- Individuals age 65 and above are included.
- Patients suffering from both hemorrhagic and ischemic stroke are included.
- Acute, sub-acute and chronic, all stages of stroke are included.

### Exclusion Criteria

- Those patients who do not want to give data by will are excluded.
- Stroke due to any accident or whiplash injury is excluded

## DATA COLLECTION PROCEDURE AND PERMISSION TAKEN FROM SUIT REASEARCH AND ETHICAL COMITTE

A relevant questionnaire wa distributed among the attendants of patients and it was filled by attendants of the patients. Data was collected through a self-modified QUESTIONNAIRE FOR VERIFYING STROKE FREE STATUS (QVSFS). After obtaining approval from the research committee, permission from director SIAHS SUIT Peshawar and from the head of the respective hospitals were taken, data were collected from the stroke, patients admitted in the public tertiary care hospitals of Peshawar who were fulfilling the eligibility criteria of study. Data were collected through self-modified QUESTIONNAIRE FOR VERIFYING STROKE FREE STATUS (QVSFS). The questionnaires were filled from the patients themselves or from/by their attendants.

## RESULTS

The main objective of this study was to find out the epidemiological status of ischemic & hemorrhagic stroke of geriatric population stoke, patients admitted in the public tertiary care hospitals of Peshawar. For this purpose, we took the data from geriatric population of age between 60 to 99 (n=109) in which the greatest percentage of stroke were in age group of 60-70 (55.0 %), followed by the age group of 71-80 (31.19 %), the age group of 81-90 (7.34 %), and the age group of 91-99(6.42 %).

### Age of Participants

| Age | Frequency | Percent |
| --- | --- | --- |
| 60-70 | 60 | 55.0 |
| 71-80 | 34 | 31.2 |
| 81-90 | 8 | 7.3 |
| 91-99 | 7 | 6.4 |
| Total | 109 | 100.0 |

Table 2 shows the gender of research participants. A total of 109 individuals were included in the study, with 59(51.13%) of the males and 50(45.87%) of females.

### Gender of research participants

| Gender | Frequency | Percent |
| --- | --- | --- |
| Male | 59 | 54.1 |
| Female | 50 | 45.9 |
| Total | 109 | 100.0 |

Table 3 shows the weights of research participants in kg. In the sample size of 109 individuals, the greatest percentage 31(28.44%) is found in the weight range of 55-65kg. Followed by 54(49.54%) is observed in the weight range of 66-75kg. Another proportion is 21(19.27%) in the 76-85kg weight range. Lastly, only 3(2.75%) in the 86-99kg weight range.

### Weight of research participants

| Weight |  | Frequency | Percent |
| --- | --- | --- | --- |
|  | 55-65 kg | 31 | 28.4 |
|  | 66-75 kg | 54 | 49.5 |
|  | 76-85 kg | 21 | 19.3 |
|  | 86-99 kg | 3 | 2.8 |
|  | Total | 109 | 100.0 |

Tables 3 shows the status of TIA/mini stroke among research participants of the 109 patients, 49 (45%) had TIA/mini stroke, whereas 60 (55%) had no history of TIA/mini stroke.

### TIA/mini stroke in research participants

|  |  | Frequency | Percent |
| --- | --- | --- | --- |
|  | Yes | 49 | 45.0 |
|  | No | 60 | 55.0 |
|  | Total | 109 | 100.0 |

### Type of Stoke

|  |  | Frequency | Percent | Valid Percent | Cumulative Percent |
| --- | --- | --- | --- | --- | --- |
|  | ischemic stoke | 78 | 71.6 | 71.6 | 71.6 |
|  | hemorrhagic stroke | 31 | 28.4 | 28.4 | 100.0 |
|  | Total | 109 | 100.0 | 100.0 |  |

Table 4 shows the type of stroke among 109 research participants, with 78 (71.56 %) having ischemic stroke and 31(28.44 %) having hemorrhagic stroke.

## DISCUSSION

According to the study conducted in tertiary care hospital in Karachi by Fawad Taj, et.al a total of 159 patients were included in this audit with 104 males (65.4%) and 55 females (34.6%). Ischemic stroke was seen in 108 patients (67.9%), while 34 patients (21.4%) with hemorrhagic stroke. Hypertension was the most prevalent risk factor present in 78% (n=124) of the population, Followed by the diabetes in 40.3% (n=64) and smoking 21% (n=33). (36). Another study conducted by Nighat Musa, et.al in Peshawar in the year of august 2018 reported that Study results showed that 62% patients were males and 38% were females. Males age distribution less than 40 years were 24% and more than 41 years were 76%. Similarly, females were 21% and 79% respectively. Major medical risk factors found were hypertension, diabetes and cardiovascular diseases. The environmental risk factors were physical inactivity, smoking and obesity. The less common risk factors were alcohol and oral contraceptive use. (8)

As our study shows the prevalence of stroke is more in males than females, a similar result were also found in the study conducted by Payam Sariaslani, et.al in Iran in the year of 2019 stating that the present study found the prevalence of stroke to be higher in men than in the women Similar results to our study regarding association of hypertension to ischemic stroke from the above study conducted by Payam Sariaslani, et.al in Iran in year 2019 they stated that The present study investigated 122 patients the most important clinical risk factors associated with ischemic stroke in the study subjects comprised a history of hypertension, heart diseases, angina and internal and rheumatic diseases, smoking, taking high-risk medications, diabetes mellitus and hyperlipidemia.(37)

## CONCLUSION

This study was performed among 109 research participants to find out the frequency of stroke in geriatric population in tertiary care hospitals of Peshawar. In current study we have concluded a high prevalence of ischemic stroke among adults aged 60 and above in tertiary care hospitals of Peshawar. The most affected were males as 51.13% and females were 45.87%.

## Data Availability

All data produced in the present work are contained in the manuscript

## References

1. Ates, S., Haarman, C. J., & Stienen, A. H. (2017). SCRIPT passive orthosis: design of interactive hand and wrist exoskeleton for rehabilitation at home after stroke. Autonomous Robots, 41(3), 711–723.

2. Banerjee, T. K., & Das, S. K. (2006). Epidemiology of stroke in India. Neurology asia, 11, 1–4.

3. Barech, M., Sadiq, S., Zarkoon, A., Gulandam, M., & Ullah, K. (2010). Risk factors for Ischemic Stroke in Patients attending Tertiary Hospital in Quetta. Pakistan Journal of Neurological Sciences (PJNS), 5(1), 1–5.

4. Center, B. (2021). Nitric Oxide-Dependent Pathways as Critical Factors in the Consequences and Recovery after Brain Ischemic Hypoxia. blood (eg, glutamate, acetylcholine, ATP), 11, 12.

5. Chaturvedi, P., Singh, A. K., Tiwari, V., & Thacker, A. K. (2020). Poststroke BDNF concentration changes following proprioceptive neuromuscular facilitation (PNF) exercises. Journal of Family Medicine and Primary Care, 9(7), 3361.

6. Chen, Z., Liu, P., Xia, X., Wang, L., & Li, X. (2022). The underlying mechanisms of cold exposure-induced ischemic stroke. Science of The Total Environment, 155514.

7. Díaz-Guzmán, J., Bermejo-Pareja, F., Benito-León, J., Vega, S., Gabriel, R., & Medrano, M. (2008). Prevalence of stroke and transient ischemic attack in three elderly populations of central Spain. Neuroepidemiology, 30(4), 247–253.

8. Eyileten, C., Jakubik, D., Shahzadi, A., Gasecka, A., van der Pol, E., De Rosa, S., … Kurkowska-Jastrzebska, I. (2022). Diagnostic Performance of Circulating miRNAs and Extracellular Vesicles in Acute Ischemic Stroke. International Journal of Molecular Sciences, 23(9), 4530.

9. Ga, D. (2008). Fisher m, macleod m, Davis Sm. Stroke. Lancet, 371(9624), 1612–1623.

10. Hanchaiphiboolkul, S., Poungvarin, N., Nidhinandana, S., Suwanwela, N. C., Puthkhao, P., Towanabut, S., … Samsen, M. (2011). Prevalence of stroke and stroke risk factors in Thailand: Thai Epidemiologic Stroke (TES) Study. Journal of the Medical Association of Thailand, 94(4), 427.

11. Hassan, K., El-Khashab, K., Mohamed, M., & Ebied, H. (2020). Left atrial size as a predictor for pulmonary hypertension in Ischemic Heart Disease. Fayoum University Medical Journal, 7(1), 88–103.

12. Hosseini, A. A., Sobhani-Rad, D., Ghandehari, K., & Benamer, H. T. (2010). Frequency and clinical patterns of stroke in Iran-Systematic and critical review. BMC neurology, 10(1), 1–10.

13. Hsieh, F.-I., & Chiou, H.-Y. (2014). Stroke: morbidity, risk factors, and care in Taiwan. Journal of stroke, 16(2), 59.

14. Huber, J., Kaczmarek, K., Leszczyńska, K., & Daroszewski, P. (2022). Post-Stroke Treatment with Neuromuscular Functional Electrostimulation of Antagonistic Muscles and Kinesiotherapy Evaluated with Electromyography and Clinical Studies in a Two-Month Follow-Up. International Journal of Environmental Research and Public Health, 19(2), 964.

15. Ikram, M. A., Wieberdink, R. G., & Koudstaal, P. J. (2012). International epidemiology of intracerebral hemorrhage. Current atherosclerosis reports, 14(4), 300–306.

16. Janssen, H., Ada, L., Middleton, S., Pollack, M., Nilsson, M., Churilov, L., … McCluskey, A. (2021). Altering the rehabilitation environment to improve stroke survivor activity: a phase II trial. International Journal of Stroke, 17474930211006999.

17. Khan, M. I., Khan, J. I., Ahmed, S. I., & Ali, S. (2019). The epidemiology of stroke in a developing country (Pakistan). Pakistan Journal of Neurological Sciences (PJNS), 13(3), 30–44.

18. Kissela, B. M., Sauerbeck, L., Woo, D., Khoury, J., Carrozzella, J., Pancioli, A., … Gebel, J. (2002). Subarachnoid hemorrhage: a preventable disease with a heritable component. Stroke, 33(5), 1321–1326.

19. Kummer, B. R., Lerario, M. P., Hunter, M. D., Wu, X., Efraim, E. S., Salehi Omran, S., … Lekic, T. (2019). Geographic analysis of mobile stroke unit treatment in a dense urban area: the new York City METRONOME registry. Journal of the American Heart Association, 8(24), e013529.

20. Kweon, S., Kim, Y., Jang, M.-j., Kim, Y., Kim, K., Choi, S., … Oh, K. (2014). Data resource profile: the Korea national health and nutrition examination survey (KNHANES). International journal of epidemiology, 43(1), 69–77.

21. Lai, J. C.-T., Wong, G. L.-H., Yip, T. C.-F., Tse, Y.-K., Lam, K. L.-Y., Lui, G. C.-Y., … Wong, V. W.-S. (2018). Chronic hepatitis B increases liver-related mortality of patients with acute hepatitis E: a territorywide cohort study from 2000 to 2016. Clinical Infectious Diseases, 67(8), 1278–1284.

22. Law, Z. K., Appleton, J. P., Scutt, P., Roberts, I., Al-Shahi Salman, R., England, T. J., … Dineen, R. A. (2022). Brief consent methods enable rapid enrollment in acute stroke trial: results from the TICH-2 randomized controlled trial. Stroke, 53(4), 1141–1148.

23. Lee, J., Jeon, J., Lee, D., Hong, J., Yu, J., & Kim, J. (2020). Effect of trunk stabilization exercise on abdominal muscle thickness, balance and gait abilities of patients with hemiplegic stroke: a randomized controlled trial. NeuroRehabilitation, 47(4), 435–442.

